# Development of Q-LAAD, an allonamer-based antigen test for the rapid detection of SARS-CoV-2

**DOI:** 10.1101/2022.09.23.22280297

**Authors:** Elise Overgaard, Shiwei Li, Hunter J. Covert, Ken Tawara, Aidan M. Poe, N. Hagan Shults, Aliona A. Chernish, Brandi Sweet, Cara R. Gonzales, Clémentine F.N. Gibard, Steven J. Burden

## Abstract

The SARS-CoV-2 virus has spread globally causing coronavirus disease 2019 (COVID-19). Rapidly and accurately identifying viral infections is an ongoing necessity. We used the systematic evolution of ligands by exponential enrichment (SELEX) technique to produce a DNA allonamer with two distinct binding domains made allosteric through a linker section; one domain binds SARS-CoV-2 spike (S) protein, inducing a conformational change that allows the reporter domain to bind a fluorescent reporter molecule. We used bead-based fluorescence and immunofluorescence assays to confirm the allonamer’s affinity and specificity for S-protein and confirmed that the allonamer can bind to S-proteins with mutations corresponding to those of the alpha, beta, gamma, and delta variants. We then developed the allonamer-based Quantum-Logic Aptamer Analyte Detection (Q-LAAD) test, a rapid, high-throughput antigen test for qualitative detection of SARS-CoV-2 in clinical settings. We validated Q-LAAD against retrospective and prospective clinical anterior nasal swab samples collected from symptomatic patients suspected of having COVID-19. Q-LAAD showed 97% sensitivity and 100% specificity compared to the RT-qPCR assay. Q-LAAD has a limit of detection (LOD) of 1.88 TCID_50_/mL, is cost-effective and convenient, and requires only a common fluorescence plate reader. Q-LAAD may be a useful clinical diagnostic tool in the fight against SARS-CoV-2.

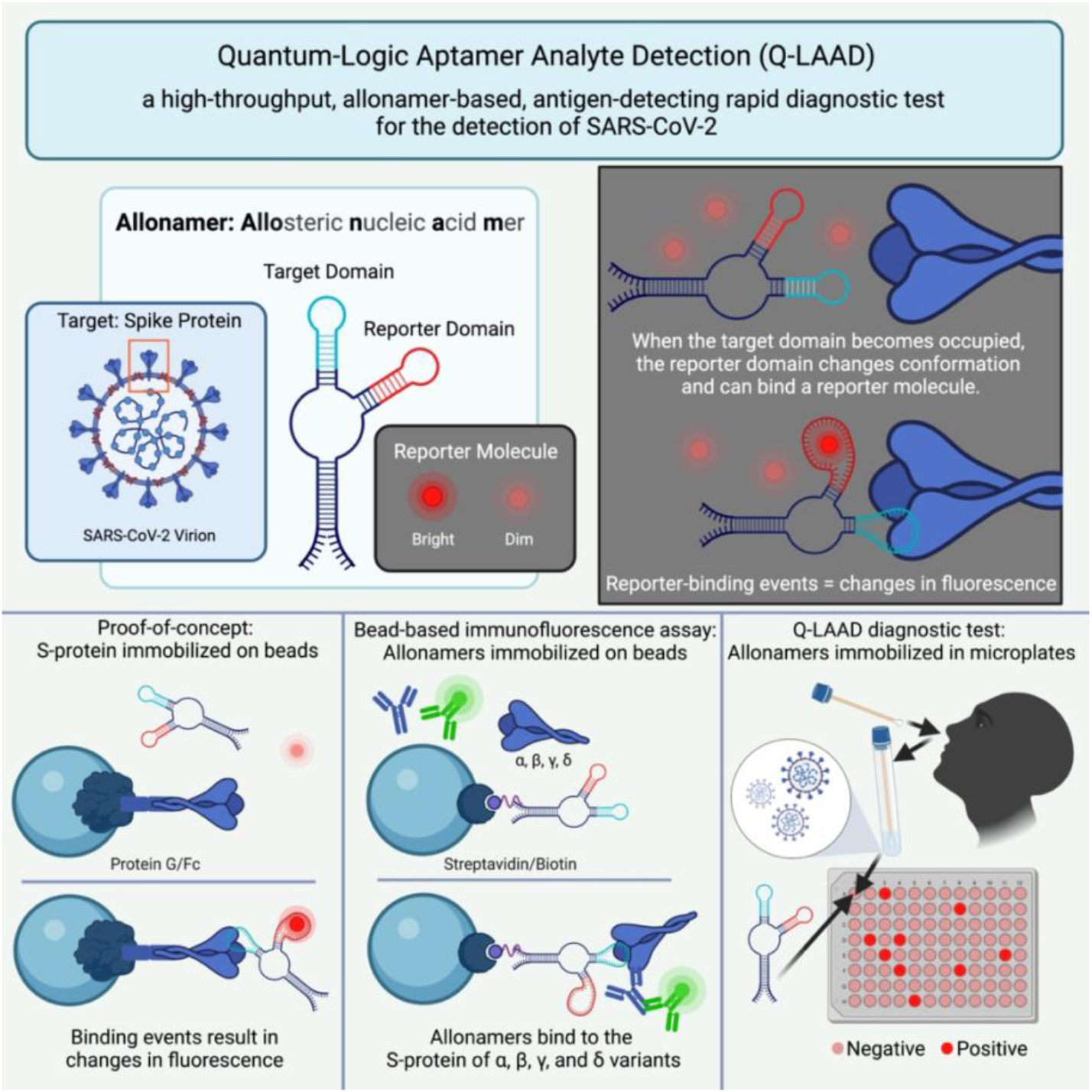

**HIGHLIGHTS:**

- Allonamers are allosterically-regulated DNA aptamers with multiple binding pockets
- Q-LAAD uses allonamers to detect SARS-CoV-2 spike protein in clinical samples
- Q-LAAD has high sensitivity and specificity and a low limit of detection
- Q-LAAD can detect spike proteins from multiple SARS-CoV-2 variants
- Q-LAAD is a dynamic, cost-effective rapid antigen test for detection of SARS-CoV-2

## 1. INTRODUCTION

The outbreak and rapid spread of the highly infectious SARS-CoV-2 virus has led to the global pandemic of coronavirus disease 2019, or COVID-19. Despite the current vaccine rollout, it is apparent that the pandemic is not ending soon. Rapid and accurate testing continues to guide public health interventions such as isolation, quarantine, and clinical management of infected individuals. Promptly and accurately identifying and isolating patients infected with SARS-CoV-2 remains a necessity.

Current methods of detection have limitations. The quantitative reverse-transcriptase polymerase chain reaction (RT-qPCR) is the current standard for detecting the SARS-CoV-2 viral genome [1–5]. SARS-CoV-2 is an enveloped, single-stranded, plus-sense RNA virus and the presence of its genome can be detected in various types of samples from the respiratory tract. RT-qPCR tests can generate false positive results, due to cross-reactions of RT-PCR primers with non-specific RNA from other viral coinfections, and/or false negative results, due to mutations occurring in the target regions of the viral genome [6]. In addition, the clinical efficiency of the RT-qPCR test is limited by laboratory capacity, appropriately trained staff, availability of expensive equipment, and the requirement for large quantities of reagent supplies [3]. Turnaround times range from several hours to several days.

The enzyme-linked immunosorbent assay (ELISA) is primarily used to measure host immune responses, mainly IgM and IgG antibody responses, to define previous exposure to SARS-CoV-2 [2–5]. Antibody tests are useful for comparing multiple samples from a single patient, but the results can vary depending on the timing of viral exposure and on the history of previous infections. In addition to having variable sensitivity and specificity, antibody tests require extensive training, have a turnaround time of several hours, and are more accurate when conducted 2 weeks after symptom onset, making them unsuitable for the purpose of rapid, early diagnosis [7,8].

SARS-CoV-2 Antigen-detecting Rapid Diagnostic Tests (Ag-RDTs) quickly detect the presence of viral proteins in the respiratory tract. Antigen presence may indicate active viral infection, which can be confirmed with an RT-qPCR test [2–5]. Two main viral proteins have been targeted for Ag-RDTs. The spike (S) protein of SARS-CoV-2 is a membrane-bound viral envelope surface protein that binds to the host cell receptor angiotensin-converting enzyme 2 (ACE2) and facilitates viral entry into the host cell. The S-protein is a heterotrimer with monomers consisting of two subunits; one subunit (S1) contains the receptor binding domain (RBD) and one subunit (S2) mediates fusion of the viral envelope and the host cell [9]. The spike (S) protein is SARS-CoV-2’s outermost surface protein, is highly immunogenic, and remains an important target for diagnosis and treatment of, and vaccination against, COVID-19 [10]. Interior to the envelope layer, the nucleocapsid (N) structural protein, both encapsulates and interacts directly with the viral genome. The N-protein is only accessible once the viral envelope is removed, so antigen tests targeting the N-protein require extra sample processing, but the N-protein is more stable and more highly conserved than the S-protein, and it is another important antigen in development of host immunity, therefore it also remains a target for viral detection [10,11]. Several companies have developed and received Emergency Use Authorization for Ag-RDTs to detect the presence of S- and N-proteins (Supplementary Table S1) [12]. In general, most of these target the N-protein and are designed as lateral flow assays, thus they are hindered by their dependence on monoclonal antibodies (mAb) that are highly specific to SARS-CoV but may not differentiate between SARS-CoV-1 and SARS-CoV-2. A small number of ELISAs are also used to detect the presence of the S- and/or N-proteins (Supplementary Table S1) [12–15].

The limitations of current testing options highlight the critical need for developing technologies that can achieve rapid, cost-effective detection of SARS-CoV-2. The challenges in developing an antigen test for SARS-CoV-2 are the ability to detect low viral loads for early identification of infection (sensitivity), to provide low or no cross-reactivity with other pathogens (specificity), and to deliver results rapidly with minimal training required [2–4].

Detecting low viral loads requires a high level of sensitivity, which can be provided by nucleic acid aptamers [16]. The use of aptamers as diagnostic tools has been considered [17–21], and the specific use of aptamers for detection of Covid-19 is currently being explored [22–29]. Aptamers are short nucleic acids with secondary structures that can form selective, high-affinity binding pockets for small molecule ligands. Aptamers can be fused to catalytic nucleic acids such as ribozymes (catalytic RNA) or deoxyribozymes (catalytic DNA) to create a ligand-triggered, catalytic molecule, or an aptazyme. Aptazymes can be developed into targeted molecular switches so that the presence of a small molecule ligand-binding event triggers a catalytic reaction. Such aptazymes are referred to as riboswitches (with catalytic RNA) or deoxyriboswitches (with catalytic DNA) and their use as diagnostic tools has been investigated for over two decades [30– 35].

Here we introduce a new class of aptamer that is similar to an aptazyme but does not employ a true catalytic mechanism. **A**llosteric **n**ucleic **a**cid **mers** (repeat units), or allonamers, are riboswitches or deoxyriboswitches that employ allosteric regulation rather than catalytic activity. During ligand binding (or unbinding) events, allonamers undergo structural rearrangements that lead to changes in the affinity of alternative binding pockets. Binding of a fluorescent molecule within an allonamer’s alternative binding pocket can influence the fluorescent output of the molecule. Like aptamers, allonamers can readily be identified by applying iterative cycles of selection and amplification to a large pool of randomized oligonucleotide sequences until a small number of catalytically active sequences with high-affinity binding pockets are discovered. We report the development and clinical validation of Quantum-Logic Aptamer Analyte Detection (Q-LAAD), a high-throughput, allonamer-based antigen test for the rapid detection of SARS-CoV-2 S-proteins in anterior nares swab specimens.

## 2. RESULTS

### 2.1. Screening and Proof-of-Concept

We used the systematic evolution of ligands by exponential enrichment (SELEX) [36,37] technique to produce a DNA allonamer with two distinct binding domains that are made allosteric through a linker section. The target domain is specific for S-protein. Binding of an S-protein to the target domain triggers restructuring of the allonamer to make the reporter domain available. A fluorescent reporter molecule can then interact with the reporter domain (see graphical abstract). Importantly, the reporter molecule can bind to the reporter domain only when the allonamer is already bound to an S-protein in the target domain. Binding of the reporter molecule in the reporter domain of the allonamer results in a change in the reporter molecule’s fluorescence. Thus, changes in the reporter molecule’s fluorescence confirm the presence of the SARS-CoV-2 S-protein.

We screened allonamers from a very large pool of oligonucleotides containing a random region for their ability to bind to the SARS-CoV-2 S1 subunit containing the RBD. After several rounds of initial selections, we used a bead-based assay to screen the remaining candidates for their selectivity for SARS-CoV-2 S1 and for their capacity to produce a fluorescent signal in the presence of the reporter molecule. Pools of candidates were incubated with recombinant Fc-tagged SARS-CoV-2 S1 immobilized on protein G-functionalized Sepharose beads, then incubated with the reporter molecule. Images of the beads showed that the pool of allonamers produced a change in fluorescent signal when incubated with SARS-CoV-2 S1. A much lower change in fluorescent signal was produced upon incubation with MERS-CoV-1 S1, indicating specificity for the SARS-CoV-2 S-protein (Figure 1).

**Figure 1:**
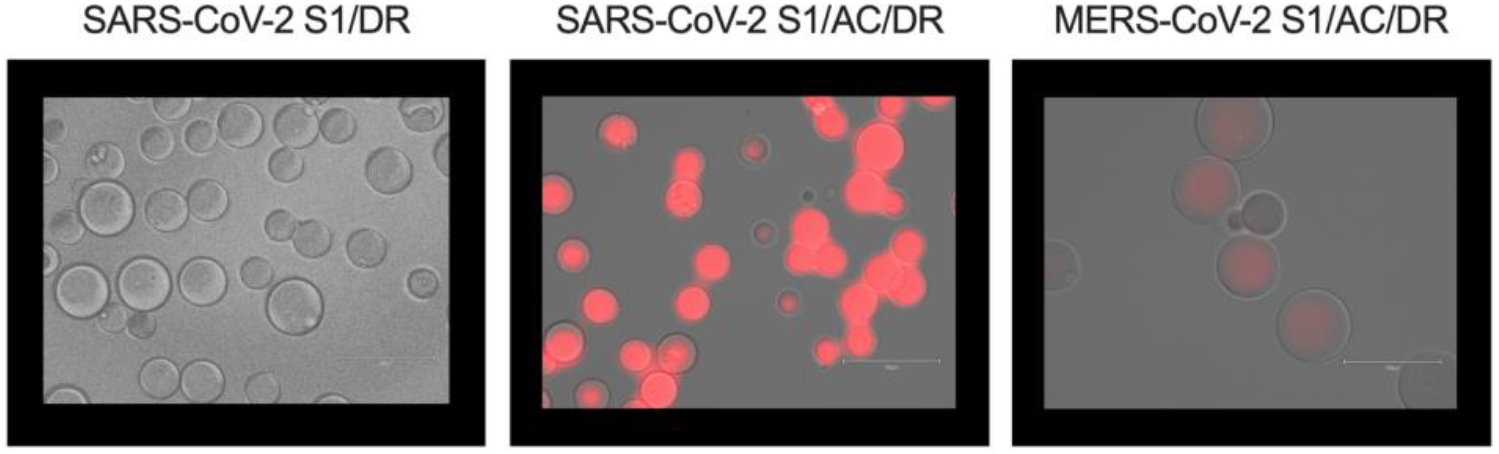
Proof-of-concept. Fc-tagged spike proteins (Fc-S1) were immobilized on protein G-functionalized Sepharose beads, then incubated with pooled allonamer candidates. Samples of incubated beads were imaged. Incubating pooled allonamer candidates with immobilized S1 proteins resulted in changes in fluorescence of the detection reagent. Pools of allonamer candidates show specificity for SARS-CoV-2 S1 compared to MERS-Cov-2 S1. Abbreviations: SARS-CoV-2 S1: immobilized SARS-CoV-2 S1 protein containing the receptor binding domain (RBD); DR: detection reagent; AC: allonamer candidate pool; MERS-CoV-2 S1: immobilized MERS-CoV-2 S1 protein containing the RBD.

After the final round of selection, the allonamer with the best performance, named N6 and part of a family in the 4-6% most abundant after sequencing, was selected for further development. Using the same bead-based assay, the allonamer still showed selectivity for SARS-CoV-2 S1 and produced a change in fluorescent signal. The N6 allonamer was further optimized through nuclease digestion-based regional mapping to reduce the total number of nucleotides. The resulting, truncated allonamer, named N6-D2, showed improved binding affinity.

### 2.2. Binding specificity studies

Our intention was to develop an assay for clinical use, which would require allonamers to be immobilized. To investigate the ability of immobilized allonamers to bind free S-protein, we immobilized 5’-biotinylated N6 or 5’-biotinylated N6-D2 allonamers on streptavidin-conjugated Sepharose beads and incubated them with free monomeric SARS-CoV-2 S1. Binding was detected with an anti-S1 RBD primary antibody and a fluorescent secondary antibody (Figure 2a). The N6 and N6-D2 allonamers produced strong signals only in the presence of free SARS-CoV-2 S1, indicating that the allonamers did bind the free S-protein (Figure 2a,b). The fluorescent signal from immobilized N6 allonamers in the presence of SARS-CoV-2 S1 was significantly higher (****p < 0.0001) than N6, N6/P/S, N6/S1/P, N6/S1/S, C/S1/P/S controls. The fluorescent signal from immobilized N6-D2 allonamers was significantly higher (****p < 0.0001) than N6 and C/S1/P/S controls, and significantly higher than N6/P/S (**p = 0.0021), N6/S1/P (**p = 0.0024), and N6/S1/S (**p = 0.0012) controls (Figure 2b).

**Figure 2:**
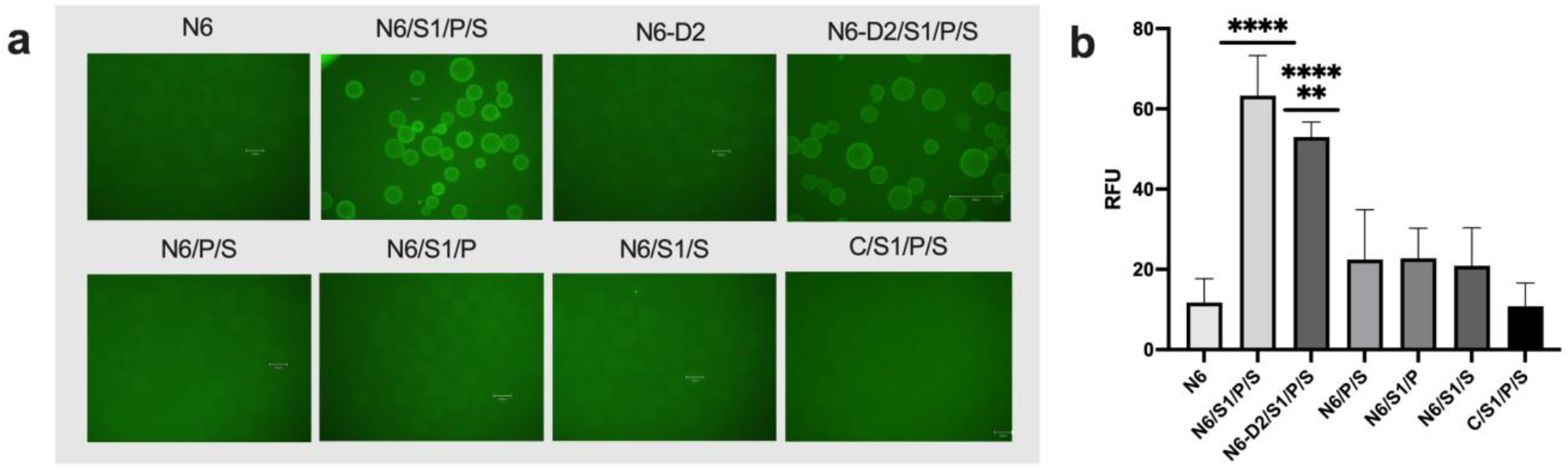
Immobilized allonamers bind free SARS-CoV-2 S1. 5’ biotinylated allonamers were immobilized on streptavidin-conjugated Sepharose beads, then incubated with free monomeric SARS-CoV-2 S1. Signals were detected with an anti-S1 receptor binding domain antibody. Samples of incubated beads were imaged. a) A fluorescent signal was detected only after N6 and N6-D2-functionalized beads were incubated with free SARS-CoV-2 S1, indicating that the immobilized allonamer did bind to free SARS-CoV-2 S1 resulting in a rearrangement of the reporter domain allowing reporter molecule to bind and causing a change in the reporter molecule’s fluorescent signal. b) Signals from experiments with immobilized N6 and N6-D2 allonamers and controls incubated with free SARS-CoV-2 S1 were quantified in relative fluorescence units (RFU). Abbreviations: N6 and N6-D2: 5’ biotinylated N6 and N6-D2 allonamers immobilized on streptavidin-conjugated Sepharose beads; P: primary antibody; S: secondary antibody; S1: SARS-CoV-2 spike protein subunit containing the receptor binding domain; C: control beads with no immobilized DNA. Data were analyzed using a one-way ANOVA with multiple comparisons. ****p < 0.0001, **p < 0.0025

We used a similar setup to investigate the specificity of the allonamer for free monomeric S1 compared to nontargets like N-protein and MERS-CoV-1 S-protein controls. Images of the beads showed a major increase in fluorescence in SARS-CoV-2 S1 compared to nontarget controls (data not shown).

Binding specificity was further investigated by incubating immobilized allonamers with various forms of SARS-CoV-2 S-protein, including free monomeric SARS-CoV-2 S1, free trimeric SARS-CoV-2 S1, heat inactivated whole SARS-CoV-2 virus, and a lentivirus expressing SARS-CoV-2 S1 (Figure 3a). Trimeric SARS-CoV-2 S1 produced a signal comparable to monomeric SARS-CoV-2 S1. The heat inactivated whole virus and lentivirus expressing SARS-CoV-2 S1 produced signals comparable to the negative control, indicating that these samples were unable to bind to the N6-D2 allonamer-functionalized beads.

**Figure 3:**
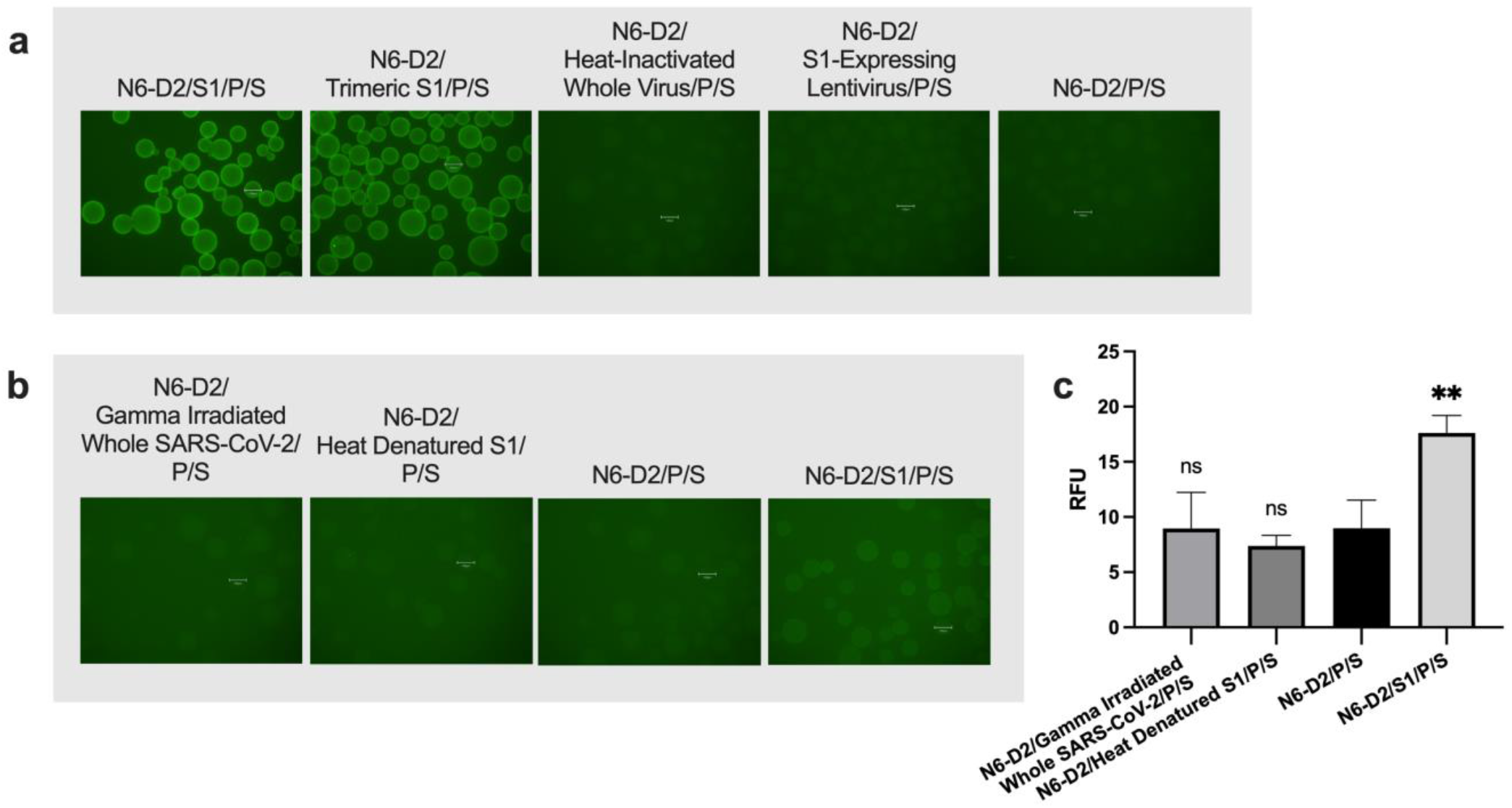
Immobilized N6-D2 allonamer binding depends on the structure of the SARS-CoV-2 S-protein. 5’ biotinylated allonamers were immobilized on streptavidin-conjugated Sepharose beads, then incubated with various forms of SARS-CoV-2 S1. Signals were detected with an anti-S1 receptor binding domain antibody. Samples of incubated beads were imaged. a) Alternative forms of the SARS-CoV-2 Spike protein tested against SARS-CoV-2 S1. b) Gamma irradiated whole SARS-CoV-2 virus and heat denatured SARS-CoV-2 S1 tested against SARS-CoV-2 S1. c) Signals from experiments with immobilized N6-D2 allonamers incubated with free gamma irradiated SARS-CoV-2 S1, heat denatured SARS-CoV-2 S1, and free SARS-CoV-2 S1 were quantified in relative fluorescence units (RFU). Abbreviations: N6-D2: 5’ biotinylated N6-D2 allonamers immobilized on streptavidin-conjugated Sepharose beads; P: primary antibody; S: secondary antibody; S1: SARS-CoV-2 spike protein subunit containing the receptor binding domain. Data were analyzed using a one-way ANOVA with multiple comparisons. **p < 0.01, ns = not significant for any comparison.

We also investigated the dependence of the allonamer on protein secondary and tertiary structure. Our intention was to develop the allonamer into a rapid antigen test for clinical samples. S-proteins may be affected by heat and/or radiation treatments, thus, dependence on S-protein structure was an important consideration in determining clinical sample handling and treatment requirements. For this study, we incubated gamma-irradiated whole SARS-CoV-2 or heat denatured SARS-CoV-2 S1 with allonamer-conjugated beads. Signal from immobilized N6-D2 allonamers in the presence of SARS-CoV-2 S1 was significantly higher (**p < 0.01) than all control groups (p = 0.0071 compared to no S1, p = 0.0070 compared to gamma irradiated whole SARS-CoV-2, p = 0.0025 compared to heat denatured S1) (Figure 3c).

Results from incubation with gamma irradiated whole SARS-CoV-2 and heat denatured SARS-CoV-2 S1 were not significant for any comparison. The N6-D2 allonamer’s inability to bind to either gamma-irradiated whole SARS-CoV-2 or heat denatured SARS-CoV-2 S1 indicates that disruption of the protein structure disrupts the ability of the protein to interact with the allonamer’s target domain. The S-protein needs to be in a native, active state, therefore clinical samples cannot be heat or radiation treated.

### 2.3. Binding affinity studies

Next, we characterized the target pocket binding affinity of the N6-D2 allonamer for SARS-CoV-2 S1 RBD (Figure 4). We incubated N6-D2 allonamer-conjugated beads with varying concentrations (0 to 400 nM) of free monomeric SARS-CoV-2 S1 (Figure 4a) and used the relative fluorescence to calculate a Kd of 22.67 nM (Figure 4b, c). We then reduced the concentration range of the free monomeric SARS-CoV-2 S1 (0 to 4 nM), repeated the assay, and used the resulting linear regression to determine the Limit of Detection (LOD) of 0.710 nM and Limit of Quantification (LOQ) of 2.558 nM according to ICH guidelines [38].

**Figure 4:**
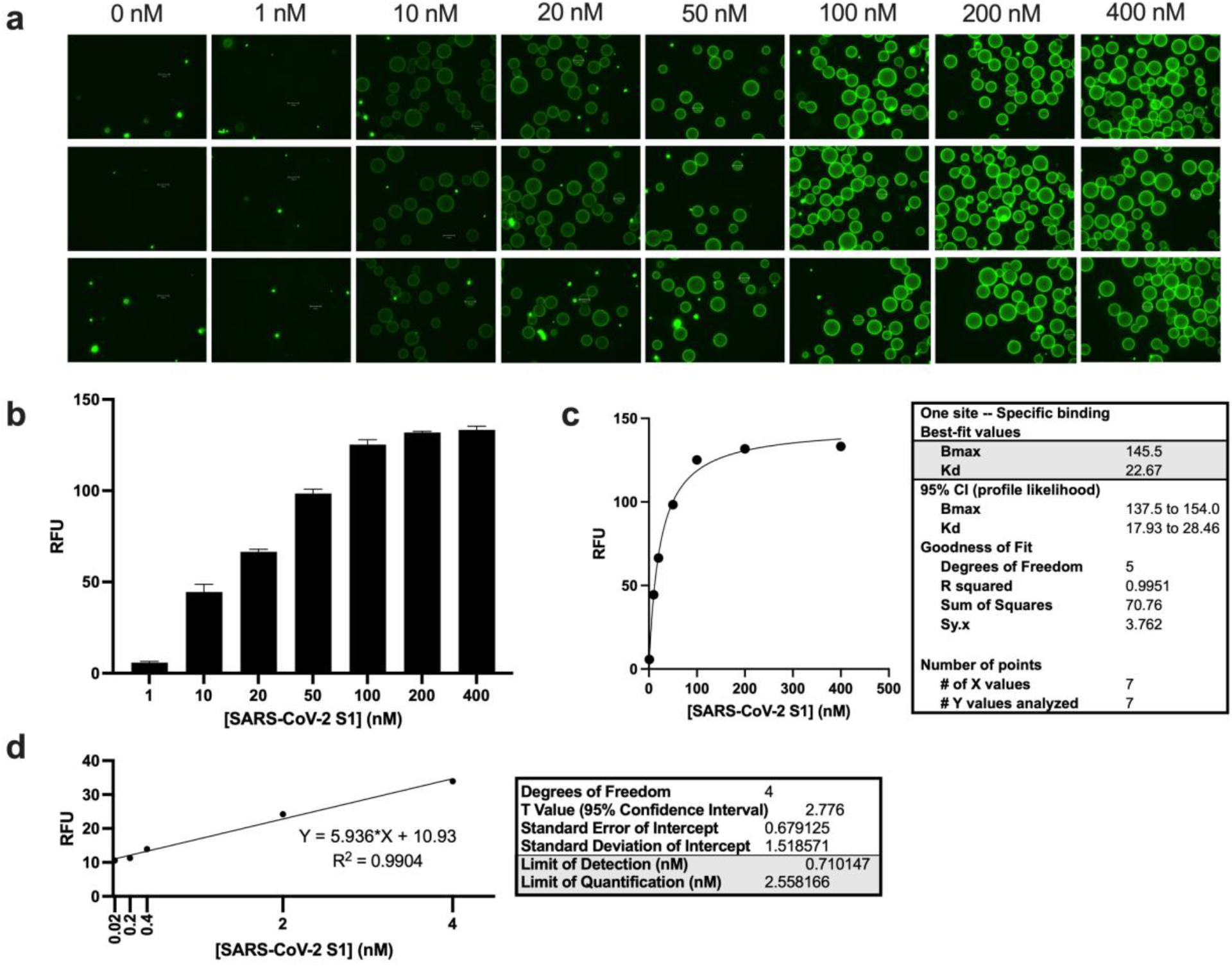
Binding affinity, Limit of Detection (LOD), and Limit of Quantification (LOQ) of immobilized allonamer. Immobilized N6-D2 aptamers were incubated with varying concentrations of SARS-CoV-2 S1. Signals were detected with an anti-S1 receptor binding domain antibody. Samples of incubated beads were imaged. a) Representative images of N6-D2 allonamer-conjugated beads incubated with varying concentrations of free SARS-CoV-2 S1. b) Signals from binding affinity experiments with immobilized N6-D2 allonamers incubated with varying concentrations of SARS-CoV-2 S1 were quantified in relative fluorescence units (RFU). c) Kd was calculated with GraphPad Prism’s one site specific binding nonlinear regression equation and found to be 22.67 nM. d) Linear regression for reduced range of SARS-CoV-2 S1 concentrations for calculations of LOD and LOQ. LOD and LOQ were calculated according to ICH guidelines [38].

### 2.4. Variant S-protein-binding studies

Some emergent SARS-CoV-2 variants carry mutations in the N- and/or S-protein genes and must be closely monitored as they may affect diagnostic tests [39–42]. Variants are classified based on their potential impact on medical countermeasures and/or prevention measures like diagnostic tests. The U.S. currently has several Variants Being Monitored (VBM, variants have a clear/potential impact, lower circulation) and Variants of Concern (VOC, variants have increased transmissibility or severity or are known to cause reduced effectiveness of treatments/vaccines or diagnostic detection failures) (Supplementary Table S2) [43].

Detecting the spike antigen despite S-protein mutations is critical to the practicality of our allonamer technology. Mutations in the S-protein could affect the ability of the S-protein to interact with our allonamer’s target domain, thus, we investigated the ability of the N6-D2 allonamer to bind S1 with mutations corresponding to those of VBM/VOC with allonamer-conjugated beads incubated with free VBM/VOC S-proteins (Figure 5). Fluorescent signals from immobilized N6 allonamers incubated with SARS-CoV-2 S1, gamma variant S1, and delta variant S1 were significantly higher (p = 0.0009, p=0.0006, p = 0.0047 respectively) than the nucleocapsid control group. Signals from incubation with alpha variant S1 and beta variant S1 showed over a five-fold change compared to the SARS-CoV-2 N-protein negative control, however, this was not significant.

**Figure 5:**
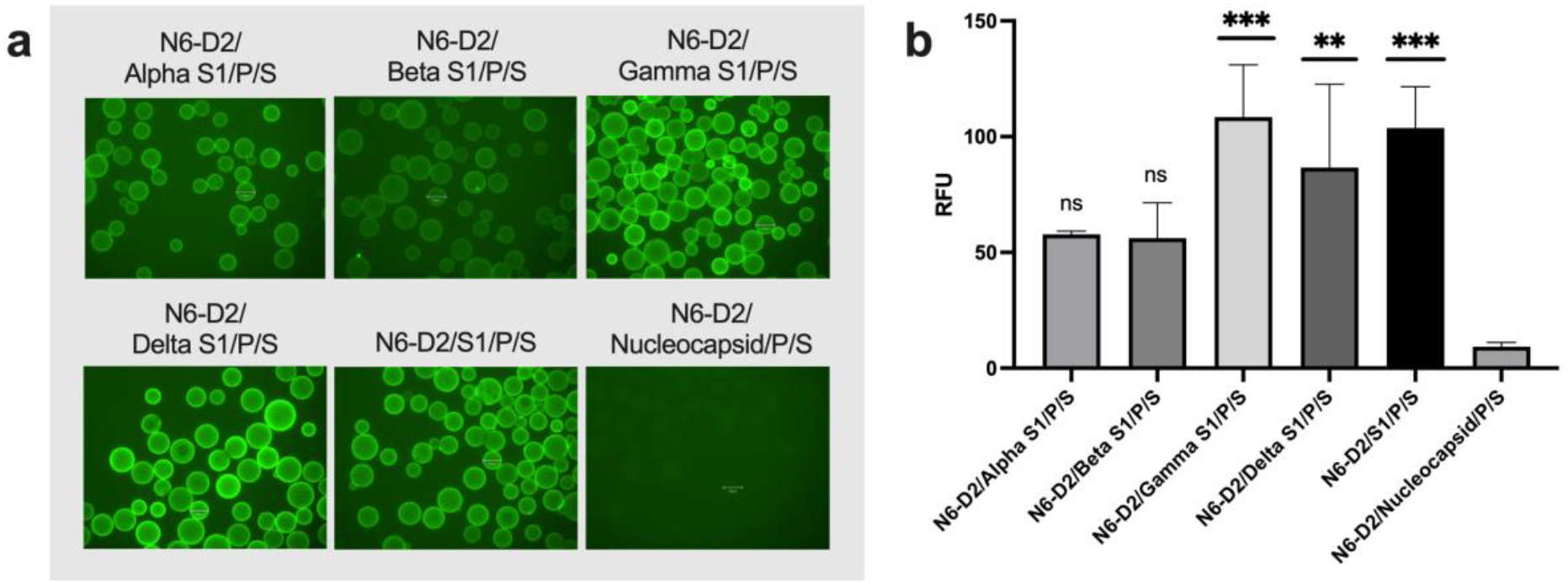
Immobilized allonamer binds recombinant SARS-CoV-2 alpha, beta, gamma, delta, and omicron variant S-proteins. Immobilized N6-D2 aptamers were incubated with SARS-CoV-2 S1 with mutations corresponding to those of current Variants Being Monitored (VBM) and Variants of Concern (VOC). Signals were detected with an anti-S1 receptor binding domain antibody. Samples of incubated beads were imaged. a) Representative images of immobilized allonamers incubated with SARS-CoV-2 S1 with mutations corresponding to those of current VBM and VOC. b) Signals from experiments with immobilized N6-D2 allonamers incubated with free wild type and variant SARS-CoV-2 S1 were quantified in relative fluorescence units (RFU). Abbreviations: Abbreviations: N6 and N6-D2: N6 and N6-D2 allonamers immobilized on beads; P: primary antibody; P2: alternate primary antibody; S: secondary antibody; S1: SARS-CoV-2 spike protein subunit containing the receptor binding domain. Data were analyzed using a one-way ANOVA with multiple comparisons. ***p < 0.001, **p < 0.01, ns = not significant

### 2.5. Clinical studies

With strong evidence from our proof-of-concept, binding specificity and affinity, and variant S-protein-binding experiments, and knowing that the change in fluorescence of the reporter molecule can be detected with a standard fluorescent plate reader, we developed Q-LAAD, a high-throughput allonamer-based Antigen-detecting Rapid Diagnostic Test (Ag-RDT) for the qualitative detection of SARS-CoV-2 in clinical anterior nares swab specimens. The Q-LAAD test is intended to be used as a rapid, high throughput test in moderate-to high-complexity laboratory settings. Q-LAAD test kits contain allonamer-functionalized (through streptavidin-biotin chemistry) microplates with the capacity to run 29 clinical samples in triplicate at once, salt-based buffer, detection reagent (with reporter molecule in solution), and positive and negative quality control solutions. As with the immobilized allonamer bead-based assays described above, allonamers immobilized in 96-well plates produce a change in the reporter molecule’s fluorescence upon reporter-binding events (see graphical abstract).

We evaluated the clinical performance of the Q-LAAD kits using both prospective (5 samples) and retrospective (60 samples) anterior nasal swab samples (total of 65 samples) from symptomatic patients suspected of having COVID-19. We compared Q-LAAD results to the results of a high-sensitivity RT-qPCR comparator method with current Emergency Use Authorization status and found the Q-LAAD tests to have very high sensitivity (97%) and specificity (100%), as well as a positive predictive value (PPV) of 100% and a negative predictive value (NPV) of 97% (Figure 6a).

**Figure 6:**
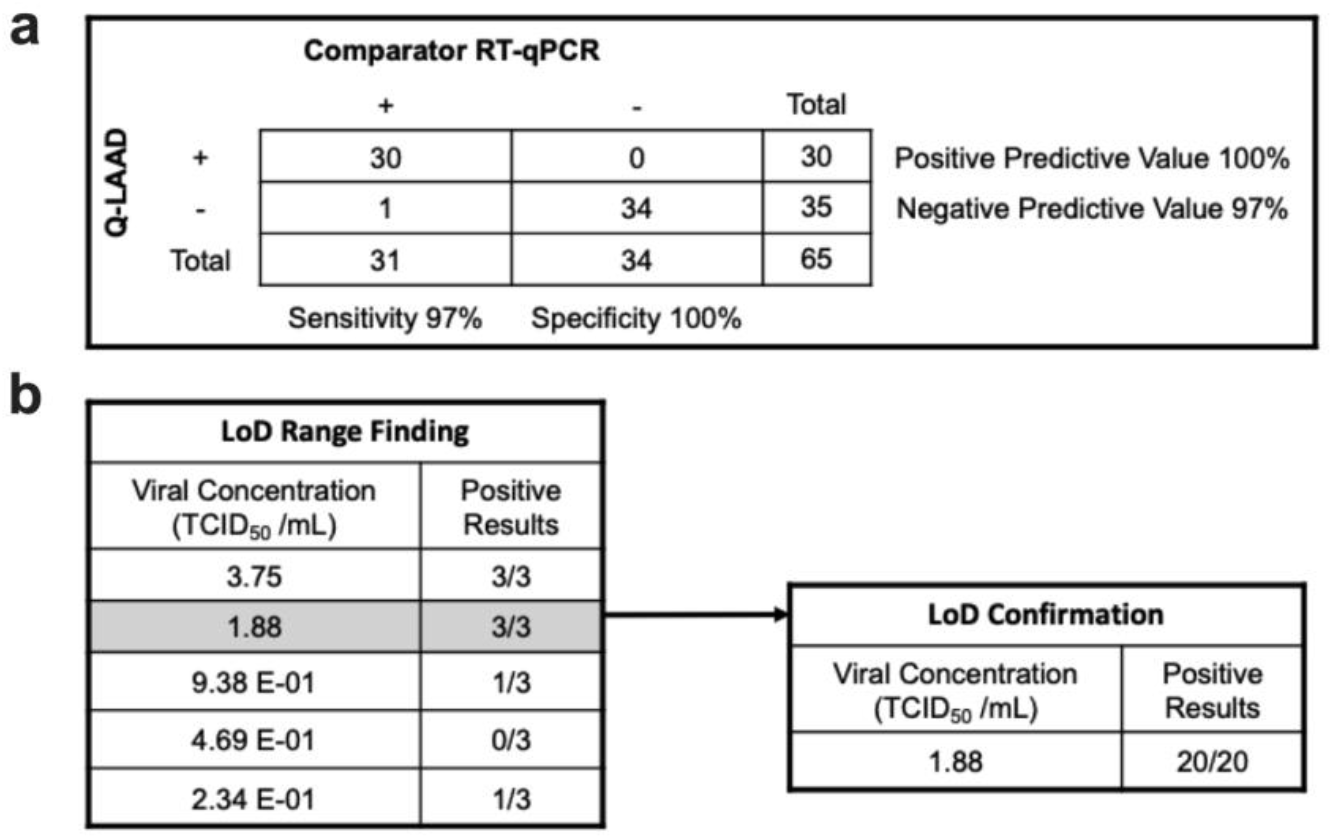
Evaluation of Q-LAAD clinical performance using both prospective and retrospective clinical samples. a) Contingency chart for calculation of the sensitivity, specificity, positive predictive value, and negative predictive value of the Q-LAAD test. b) Limiting dilution study using positive clinical specimens for calculation of Limit of Detection (LOD). LOD was confirmed using positive clinical specimens.

We also used the clinical samples to determine the limit of detection (LoD), or the lowest detectable concentration of SARS-CoV-2 at which approximately 95% of all true positive replicates test positive. The LoD was determined with limiting dilution studies using confirmed positive clinical specimens and was found to be 1.88 TCID_50_/mL (Figure 6b).

It is also important to consider factors that could potentially cross-react or interfere with the Q-LAAD assay. We evaluated the cross-reactivity of the assay with 28 high-priority pathogens that are commonly found in the respiratory tract flora. We observed no cross-reactivity (Supplementary Table S3) or test interference (Supplementary Table S4) from any of the tested microbes. We also evaluated the effects of 19 endogenous substance that could potentially interfere with the assay. None of the tested endogenous substances interfered with the detection of SARS-CoV-2 (Supplementary Table S5).

## 3. DISCUSSION

Our studies showed that allonamer technology can be used to detect the presence of SARS-CoV-2 in clinical anterior nares swab samples in *in vitro* assays. Q-LAAD combines the speed and relative simplicity of an antigen test with the sensitivity of a molecular test. With a binding affinity for SARS-CoV-2 S-proteins in the low nanomolar range, a limit of detection lower than most comparator technologies, and no cross-reactivity with or interference from high-priority pathogens commonly found in the respiratory tract flora, the allonamer-based Q-LAAD technology could be very valuable in the fight against SARS-CoV-2. The sensitivity (97%, indicating the probability of correctly identifying patients with SARS-CoV-2 infection) and specificity (100%, indicating the probability of correctly identifying patients with no SARS-CoV-2 infection) of Q-LAAD in comparison to the current standard exceed the World Health Organization’s minimum performance requirements (≥ 80% sensitivity and ≥ 97% specificity) for Ag-RDT’s to be recommended for use [44]. The positive predictive value indicates that if the Q-LAAD test is positive, the probability of SARS-CoV-2 infection is 100%. The negative predictive value indicates that if the Q-LAAD test is negative, the probability of not having a SARS-CoV-2 infection is 97%. These metrics show that Q-LAAD should be highly competitive as a SARS-CoV-2 diagnostic test.

We were unable to confirm binding of bead-immobilized allonamers to whole inactivated virus and/or lentivirus expressing SARS-CoV-2 S1. This is consistent with findings from previous internal studies where we found that positive clinical samples were unable to bind to bead-immobilized allonamers. It is possible that the larger targets were unable to bind due to steric hindrance between the beads and the large viral complexes. In addition, the heating process for the whole heat-inactivated viral samples may have affected the structure of the S-protein. Regardless, the bead-immobilized assays were only performed to provide proof-of-concept evidence. We did show that clinical samples were able to bind to allonamers immobilized on microplates, therefore the issues with bead-immobilized allonamers are irrelevant to the final Q-LAAD assay product.

Although no endogenous substances interfered with the assay, some did show cross-reactive activity. Blood is a well-established inhibitor of fluorescence. Hemoglobin has been shown to be a potent quencher of free dyes such as EvaGreen, rhodamine based dye, and 6-carboxy-X-rhodamine [45]. Q-LAAD utilizes free rhodamine dye molecules in the detection reagent. To mitigate false positives, samples suspected of having blood present should be recollected or tested using a different method. We do not report any false negatives due to the presence of blood up to 4% v/v. Other cross-reactive substances included purified mucin protein, mupirocin, phenylephrine hydrochloride, oseltamivir, and oxymetazoline hydrochloride. Purified mucin proteins were cross-reactive at 0.5% w/v, which means the clinical accuracy of Q-LAAD is reduced if there are large amounts of mucus on the collection swab and efforts should be made during collection to minimize accumulation of mucus. The remaining cross-reactive substances are ingredients in commonly used treatments for upper respiratory tract infections. To avoid cross-reactivity in the assay, we recommend that samples be collected after appropriate washouts from these treatments.

## 4. CONCLUSIONS

The Q-LAAD assay requires small amounts of reagents that are readily available, even during the current pandemic, and we showed that the allonamers do interact with current important Variants of Concern/Variants Being Monitored. Should a variant emerge that our allonamer cannot detect, the ease and simplicity of the SELEX process ensures that allonamers may be updated/replaced quickly and efficiently if necessary. We chose to develop an assay that targeted the S-protein. Targeting the outermost surface protein requires less time and equipment and fewer reagents for sample processing (the envelope must be removed to access N-proteins), however, the flexibility of the allonamer technology also means that N-proteins could be easily incorporated into a more robust, multiple-target assay in the future. Finally, the 96-well plate format is easy to set up and it requires only five minutes of reading time on a BioTek plate reader with Gen5 software, a common plate reader in moderate-complexity labs. With an optimized workflow, the Q-LAAD assay can yield a throughput of 348 tests per hour per instrument. As the SARS-CoV-2 pandemic carries on, the Q-LAAD assay will be a dynamic and useful tool for early detection of infection.

## 5. MATERIALS AND METHODS

**Table 1:** Key Resources.

| Reagent/Resource | Source | Identifier |
| --- | --- | --- |
| SARS-CoV-2 S1-Fc | The Native Antigen Company | REC31806-100 |
| MERS-CoV-1 S1-Fc | The Native Antigen Company | REC31847-100 |
| SARS-CoV-2 monomeric S1-His | Sino Biological | 40591-V08H |
| SARS-CoV-2 trimeric S1-His | The Native Antigen Company | REC31871-100 |
| SARS-CoV-2 alpha S1-His-Strep | The Native Antigen Company | REC31924-100 |
| SARS-CoV-2 beta S1-His-Strep | The Native Antigen Company | REC31926-100 |
| SARS-CoV-2 gamma S1-His | Sino Biological | 40592-V08H86 |
| SARS-CoV-2 delta S1 RBD-Fc | Sino Biological | 40592-V02H3 |
| SARS-CoV-2 N-protein-His | The Native Antigen Company | REC31812-100 |
| Whole heat-inactivated SARS-CoV-2 | Helix Elite™ | HE0065N |
| Whole gamma-irradiated SARS-CoV-2 | bei Resources | NR-52287 |
| Lentivirus expressing SARS-CoV-2 S1 | Gifted: University of Manitoba | N/A |
| Protein G Sepharose Beads | BioVision | 6511 |
| Streptavidin-Sepharose Beads | BioVision | 6565-2 |
| Extra Sense™ BCA Protein Assay Kit | BioVision | K814 |
| Rabbit Anti-SARS-CoV-2 S1 RBD | Absolute Antibody | Ab02019-23.0 [CV30] |
| Donkey Anti-Rabbit IgG NL493-Conjugated | R&D Systems | NL006 |

### 5.1. Buffers and chemicals

Dulbecco’s Phosphate Buffered Saline (DPBS) was purchased from Cytiva/Hyclone Laboratories. Bovine Serum Albumin (BSA) was purchased from Fisher BioReagents. TWEEN^®^ 20, sodium chloride, Trizma^®^ base, magnesium chloride hexahydrate, and potassium chloride were purchased from Sigma Aldrich.

### 5.2. SELEX

SELEX was performed as previously described [36,37]. Sequence data patent pending.

### 5.3. Fc-S1 bead-based proof-of-concept assay

Fc-tagged SARS-CoV-2 S1 or Fc-tagged MERS-CoV-2 S1 was immobilized on protein G-functionalized Sepharose beads. Bicinchoninic acid assays (BCAs) were performed before and after conjugation to quantify protein and confirm conjugation. S1-Fc-conjugated beads were incubated at room temperature with either the pool of allonamer candidates or individual, unique allonamers for 5 minutes, then rinsed 3 times with bead washing buffer (BWB, 100mM NaCl, 20mM Tris, 5mM MgCl2, 2mM KCl in nanopure water) to ensure removal of non-specific allonamers. Rinsed beads with bound allonamers were incubated at room temperature with the reporter molecule for 3 minutes, then rinsed 3 times with BWB to remove excess reporter molecule. Beads were analyzed with an EVOS M5000 imaging system with a GFP filter. A high signal-to-noise ratio was used to allow for detection of the fluorescent signal emitted by the reporter molecule.

### 5.4. Immobilized allonamer target-binding assays

5’-biotinylated allonamers were immobilized on streptavidin-conjugated Sepharose beads. Un-conjugated allonamers were removed with BWB wash steps. Immobilized DNA was quantified using a Qubit Fluorometer (Invitrogen).

Immobilized allonamers were then incubated with free SARS-CoV-2 S1-His, SARS-CoV-2 trimeric S1-His, whole heat-inactivated SARS-CoV-2, lentivirus expressing SARS-CoV-2 S1, or SARS-CoV-2 S1 with Variants of Concern/Variants Being Monitored mutations. Unbound targets were removed with BWB wash steps Binding was detected with a rabbit anti-SARS-CoV-2 S1 RBD primary antibody and a fluorescent donkey anti-rabbit IgG secondary antibody. Fluorescence was detected and analyzed using an EVOS M5000 imaging system with a GFP filter. All target proteins and primary and secondary antibody dilutions were made in phosphate-buffered saline + TWEEN^®^ 20 (PBST) buffer (1% BSA, 0.1% TWEEN^®^ 20, 100 nM NaCl, 20 mM Tris, 5 mM MgCl_2_, 2 mM KCl in DPBS).

For heat-inactivation and gamma-irradiation studies, SARS-CoV-2 S1-His was incubated at 65°C for 30 minutes prior to use. Gamma-irradiated whole virus was purchased (see Key Resources table). Immobilized allonamer target-binding assays were completed as described above.

For immobilized allonamer target-binding experiments, assays were performed in biological quadruplicates, and for each biological replicate, three samples were collected and used for imaging. One representative image from each biological replicate was selected for data analysis. Signals were quantified in relative fluorescence units with background (buffer only) subtracted. Data shown is an average of four samples, one from each biological replicate.

For S-protein form dependence experiments, assays were performed in biological triplicates, and for each biological replicate, three samples were collected and used for imaging. One representative image from each biological replicate was selected for data analysis. Signals were quantified in relative fluorescence units with background (buffer only) subtracted. Data shown is an average of three samples, one from each biological replicate.

For variant experiments, assays were performed in biological triplicates, and for each biological replicate, three samples were collected and used for imaging. Signals from all three images were quantified in relative fluorescence units with background (buffer only) subtracted. Data shown includes three quantified samples from three biological replicates.

### 5.5. Calculation of Kd, Limit of Detection (LOD), and Limit of Quantification (LOQ)

N6-D2 allonamers were immobilized, washed, and quantified as described above, then incubated with varying concentrations of SARS-CoV-2 S1. Assays were performed in biological triplicates, and for each biological replicate, three samples were collected and used for imaging. Signals were quantified in relative fluorescence units with background (buffer only) subtracted. Data shown is an average of triplicate samples from one representative biological replicate. Kd was calculated with GraphPad Prism using the One Site Specific Binding nonlinear regression equation. Limit of Detection (LOD) and Limit of Quantification (LOQ) were calculated according to the International Council on Harmonization (ICH) Tripartite Guideline - Validation Of Analytical Procedures: Text And Methodology Q2(R1) document’s guidelines for determining detection and quantitation limits. [38]

### 5.6. Cross-reactivity and interference assays

Microbes, pools of microbes, or endogenous substances were included in the Q-LAAD assay at the concentrations noted in Supplementary Tables 3-5. Low viral-load positive samples were used at 3x LoD.

### 5.7. Clinical study

Retrospective clinical samples were collected with IRB approval from third-party vendors at different locations across the United States. Deidentified samples were eluted in 0.9% saline buffer, shipped frozen, thawed and aliquoted upon receipt, then stored at −80°C. Samples underwent no more than two freeze-thaw cycles in total. Prospective clinical samples were collected with IRB approval and with written informed consent. Deidentified samples were eluted in 0.9% saline buffer, aliquoted, then stored at −80°C until tested. All samples were randomized and tested for presence of SARS-CoV-2 using the Q-LAAD high throughput kit. Presence of SARS-CoV-2 in clinical samples was verified using a high-sensitivity RT-qPCR comparator method with current Emergency Use Authorization status. Q-LAAD testing was conducted by operators blinded to the reference RT-qPCR result.

## Data Availability

All data produced in the present study are available upon reasonable request to the authors.

## 6. LIST OF SUPPLEMENTARY MATERIALS

Table S1: SARS-CoV-2 Antigen Diagnostic Tests with Emergency Use Authorization

Table S2: SARS-CoV-2 Variants and Current SARS-CoV-2 Interagency Group (SIG) Classification Table S3: Cross-Reactivity of High-Priority Pathogens Commonly Found in the Respiratory Tract Flora Table S4: Interference of High-Priority Pathogens Commonly Found in the Respiratory Tract Flora

Table S5: Interference of Endogenous Substances

## 7. ACKNOWLEDGEMENTS

Graphical abstract and graphical representations of experimental setup for figures were created with BioRender.com.

## 8. AUTHOR CONTRIBUTIONS

Authors confirm contribution to the paper as follows: **Elise Overgaard**: Data curation, Visualization, Writing - original draft, Writing - review & editing; **Shiwei Li**: Conceptualization, Data curation, Formal analysis, Investigation, Methodology, Project administration, Supervision, Validation, Visualization, Writing - review & editing; **Hunter J. Covert:** Conceptualization, Data curation, Formal analysis, Investigation, Methodology, Project administration, Supervision, Validation, Visualization, Writing - review & editing; **Ken Tawara:** Data curation, Investigation, Software, Writing - review & editing; **Aidan M. Poe:** Investigation, Writing - review & editing; **N. Hagan Shults:** Investigation, Writing - review & editing; **Aliona A. Chernish:** Investigation, Writing - review & editing; **Brandi Sweet:** Investigation, Writing - review & editing; **Cara R. Gonzales:** Investigation, Writing - review & editing; **Clémentine F.N. Gibard:** Conceptualization, Data curation, Formal analysis, Funding acquisition, Investigation, Methodology, Project administration, Supervision, Validation, Visualization, Writing - review & editing; **Steven J. Burden:** Conceptualization, Data curation, Formal analysis, Funding acquisition, Investigation, Methodology, Project administration, Supervision, Validation, Visualization, Writing - review & editing.

All authors reviewed the results and approved the final version of the manuscript.

## REFERENCES

[1] R. Pu, S. Liu, X. Ren, D. Shi, Y. Ba, Y. Huo, W. Zhang, L. Ma, Y. Liu, Y. Yang, N. Cheng, The screening value of RT-LAMP and RT-PCR in the diagnosis of COVID-19: systematic review and meta-analysis, J. Virol. Methods. 300 (300) 114392. https://doi.org/10.1016/J.JVIROMET.2021.114392.

[2] P. Rai, B.K. Kumar, V.K. Deekshit, I. Karunasagar, I. Karunasagar, Detection technologies and recent developments in the diagnosis of COVID-19 infection, Appl. Microbiol. Biotechnol. 105 (105) 441–455. https://doi.org/10.1007/s00253-020-11061-5.

[3] L.J. Carter, L. V. Garner, J.W. Smoot, Y. Li, Q. Zhou, C.J. Saveson, J.M. Sasso, A.C. Gregg, D.J. Soares, T.R. Beskid, S.R. Jervey, C. Liu, Assay Techniques and Test Development for COVID-19 Diagnosis, ACS Cent. Sci. 6 (6) 591–605. https://doi.org/10.1021/ACSCENTSCI.0C00501/SUPPL_FILE/OC0C00501_SI_001.PDF.

[4] B.D. Kevadiya, J. Machhi, J. Herskovitz, M.D. Oleynikov, W.R. Blomberg, N. Bajwa, D. Soni, S. Das, M. Hasan, M. Patel, A.M. Senan, S. Gorantla, J.E. McMillan, B. Edagwa, R. Eisenberg, C.B. Gurumurthy, S.P.M. Reid, C. Punyadeera, L. Chang, H.E. Gendelman, Diagnostics for SARS-CoV-2 infections, Nat. Mater. 2021 205. 20 (20) 593–605. https://doi.org/10.1038/s41563-020-00906-z.

[5] M. Yüce, E. Filiztekin, K.G. Özkaya, COVID-19 diagnosis —A review of current methods, Biosens. Bioelectron. 172 (172) 112752. https://doi.org/10.1016/J.BIOS.2020.112752.

[6] A. Tahamtan, A. Ardebili, Real-time RT-PCR in COVID-19 detection: issues affecting the results, Https://Doi.Org/10.1080/14737159.2020.1757437. 20 (20) p453–454. https://doi.org/10.1080/14737159.2020.1757437.

[7] M. Chen, R. Qin, M. Jiang, Z. Yang, W. Wen, J. Li, Clinical applications of detecting IgG, IgM or IgA antibody for the diagnosis of COVID-19: A meta-analysis and systematic review, Int. J. Infect. Dis. 104 (104) 415–422. https://doi.org/10.1016/J.IJID.2021.01.016.

[8] J. Kopel, H. Goyal, A. Perisetti, Antibody tests for COVID-19, Baylor Univ. Med. Cent. Proc. 34 (34) 63–72. https://doi.org/10.1080/08998280.2020.1829261.

[9] D. Wrapp, N. Wang, K.S. Corbett, J.A. Goldsmith, C.-L. Hsieh, O. Abiona, B.S. Graham, J.S. Mclellan, Cryo-EM structure of the 2019-nCoV spike in the prefusion conformation, 2019. http://science.sciencemag.org/ (accessed February 3, 2021).

[10] S. Soleimanpour, A. Yaghoubi, COVID-19 vaccine: where are we now and where should we go?, Https://Doi.Org/10.1080/14760584.2021.1875824. 20 (20) p23–44. https://doi.org/10.1080/14760584.2021.1875824.

[11] W. Zeng, G. Liu, H. Ma, D. Zhao, Y. Yang, M. Liu, A. Mohammed, C. Zhao, Y. Yang, J. Xie, C. Ding, X. Ma, J. Weng, Y. Gao, H. He, T. Jin, Biochemical characterization of SARS-CoV-2 nucleocapsid protein, Biochem. Biophys. Res. Commun. 527 (527) 618–623. https://doi.org/10.1016/J.BBRC.2020.04.136.

[12] U.S. Food and Drug Administration, In Vitro Diagnostics EUAs - Antigen Diagnostic Tests for SARS-CoV-2 | FDA, (2021) 1–5. https://www.fda.gov/medical-devices/coronavirus-disease-2019-covid-19-emergency-use-authorizations-medical-devices/in-vitro-diagnostics-euas-antigen-diagnostic-tests-sars-cov-2 (accessed February 19, 2022).

[13] M.M. Żak, A. Stock, D. Stadlbauer, W. Zhang, K. Cummings, W. Marsiglia, A. Zargarov, F. Amanat, M. Tamayo, C. Cordon-Cardo, F. Krammer, D.R. Mendu, Development and characterization of a quantitative ELISA to detect anti-SARS-CoV-2 spike antibodies, Heliyon. 7 (7) e08444. https://doi.org/10.1016/J.HELIYON.2021.E08444.

[14] V. Krähling, S. Halwe, C. Rohde, D. Becker, S. Berghöfer, C. Dahlke, M. Eickmann, M.S. Ercanoglu, L. Gieselmann, A. Herwig, A. Kupke, H. Müller, P. Neubauer-Rädel, F. Klein, C. Keller, S. Becker, Development and characterization of an indirect ELISA to detect SARS-CoV-2 spike protein-specific antibodies, J. Immunol. Methods. 490 (490) 112958. https://doi.org/10.1016/J.JIM.2021.112958.

[15] E.S. Theel, J. Harring, H. Hilgart, D. Granger, Performance characteristics of four high-throughput immunoassays for detection of igg antibodies against sars-cov-2, J. Clin. Microbiol. 58 (2020). https://doi.org/10.1128/JCM.01243-20/ASSET/FF7749D0-9137-40D0-A46D-08786E2B8C69/ASSETS/GRAPHIC/JCM.01243-20-F0002.JPEG.

[16] A. Krüger, A.P. de Jesus Santos, V. de Sá, H. Ulrich, C. Wrenger, Aptamer Applications in Emerging Viral Diseases, Pharm. 2021, Vol. 14, Page 622. 14 (14) 622. https://doi.org/10.3390/PH14070622.

[17] P.K. Kulabhusan, B. Hussain, M. Yüce, Current Perspectives on Aptamers as Diagnostic Tools and Therapeutic Agents, Pharmaceutics. 12 (12) 1–23. https://doi.org/10.3390/PHARMACEUTICS12070646.

[18] G.T. Rozenblum, V.G. Lopez, A.D. Vitullo, M. Radrizzani, Aptamers: current challenges and future prospects, Expert Opin. Drug Discov. 11 (11) 127–135. https://doi.org/10.1517/17460441.2016.1126244.

[19] A. V. Lakhin, V.Z. Tarantul, L. V. Gening, Aptamers: Problems, Solutions and Prospects, Acta Naturae. 5 (5) 34. https://doi.org/10.32607/20758251-2013-5-4-34-43.

[20] E. Iturriaga-Goyon, B. Buentello-Volante, F.S. Magaña-Guerrero, Y. Garfias, Future Perspectives of Therapeutic, Diagnostic and Prognostic Aptamers in Eye Pathological Angiogenesis, Cells. 10 (2021). https://doi.org/10.3390/CELLS10061455.

[21] O.O. Ayodele, A.O. Adesina, S. Pourianejad, J. Averitt, T. Ignatova, Recent Advances in Nanomaterial-Based Aptasensors in Medical Diagnosis and Therapy, Nanomaterials. 11 (2021). https://doi.org/10.3390/NANO11040932.

[22] J. Byun, Recent Progress and Opportunities for Nucleic Acid Aptamers, Life (Basel, Switzerland). 11 (11) 1–18. https://doi.org/10.3390/LIFE11030193.

[23] L. Zhang, X. Fang, X. Liu, H. Ou, H. Zhang, J. Wang, Q. Li, H. Cheng, W. Zhang, Z. Luo, Discovery of sandwich type COVID-19 nucleocapsid protein DNA aptamers, Chem. Commun. 56 (56) 10235–10238. https://doi.org/10.1039/D0CC03993D.

[24] C. Acquah, J. Jeevanandam, K.X. Tan, M.K. Danquah, Engineered Aptamers for Enhanced COVID-19 Theranostics, Cell. Mol. Bioeng. 14 (14) 209–221. https://doi.org/10.1007/S12195-020-00664-7/FIGURES/4.

[25] S. Krishnan, A. Kumar Narasimhan, D. Gangodkar, S. Dhanasekaran, N. Kumar Jha, K. Dua, V.K. Thakur, P. Kumar Gupta, Aptameric nanobiosensors for the diagnosis of COVID-19: An update, Mater. Lett. 308 (308) 131237. https://doi.org/10.1016/J.MATLET.2021.131237.

[26] L. Shi, L. Wang, X. Ma, X. Fang, L. Xiang, Y. Yi, J. Li, Z. Luo, G. Li, Aptamer-Functionalized Nanochannels for One-Step Detection of SARS-CoV-2 in Samples from COVID-19 Patients, Anal. Chem. 93 (93) 16646–16654. https://doi.org/10.1021/ACS.ANALCHEM.1C04156/SUPPL_FILE/AC1C04156_SI_001.PDF.

[27] J. Tian, Z. Liang, O. Hu, Q. He, D. Sun, Z. Chen, An electrochemical dual-aptamer biosensor based on metal-organic frameworks MIL-53 decorated with Au@Pt nanoparticles and enzymes for detection of COVID-19 nucleocapsid protein, Electrochim. Acta. 387 (387) 138553. https://doi.org/10.1016/J.ELECTACTA.2021.138553.

[28] A. Gupta, A. Anand, N. Jain, S. Goswami, A. Anantharaj, S. Patil, R. Singh, A. Kumar, T. Shrivastava, S. Bhatnagar, G.R. Medigeshi, T.K. Sharma, A novel G-quadruplex aptamer-based spike trimeric antigen test for the detection of SARS-CoV-2, Mol. Ther. - Nucleic Acids. 26 (26) 321–332. https://doi.org/10.1016/J.OMTN.2021.06.014.

[29] F. Morena, C. Argentati, I. Tortorella, C. Emiliani, S. Martino, De novo ssRNA Aptamers against the SARS-CoV-2 Main Protease: In Silico Design and Molecular Dynamics Simulation, Int. J. Mol. Sci. 22 (2021). https://doi.org/10.3390/IJMS22136874.

[30] J.J. Rossi, K. Kruger, P.J. Grabowski, A.J. Zaug, Ribozyme Diagnostics Comes of Age, Chem. Biol. 11 (11) 894–895. https://doi.org/10.1016/J.CHEMBIOL.2004.07.002.

[31] A. Shih, J.M. Bockman, S.T. George, United States Patent: 5589332, 5,589,332, 1996. https://patft.uspto.gov/netacgi/nph-Parser?Sect1=PTO2&Sect2=HITOFF&u=%2Fnetahtml%2FPTO%2Fsearch-adv.htm&r=120&f=G&l=50&d=PTXT&p=3&S1=5,589,332&OS=5,589,332&RS=5,589,332 (accessed February 24, 2022).

[32] R.R. Breaker, DNA enzymes, Nat. Biotechnol. 1997 155. 15 (15) 427–431. https://doi.org/10.1038/nbt0597-427.

[33] R.R. Breaker, G.F. Joyce, The expanding view of RNA and DNA function, Chem. Biol. 21 (21) 1059–1065. https://doi.org/10.1016/j.chembiol.2014.07.008.

[34] H. Wei, B. Li, J. Li, S. Dong, E. Wang, DNAzyme-based colorimetric sensing of lead (Pb(2+)) using unmodified gold nanoparticle probes, Nanotechnology. 19 (2008). https://doi.org/10.1088/0957-4484/19/9/095501.

[35] T. Lan, K. Furuya, Y. Lu, A highly selective lead sensor based on a classic lead DNAzyme, Chem. Commun. 46 (46) 3896–3898. https://doi.org/10.1039/B926910J.

[36] C. Chai, Z. Xie, E. Grotewold, SELEX (Systematic Evolution of Ligands by EXponential Enrichment), as a powerful tool for deciphering the protein-DNA interaction space, Methods Mol. Biol. 754 (754) 249–258. https://doi.org/10.1007/978-1-61779-154-3_14.

[37] F. Spill, Z.B. Weinstein, A.I. Shemirani, N. Ho, D. Desai, M.H. Zaman, Controlling uncertainty in aptamer selection, Proc. Natl. Acad. Sci. U. S. A. 113 (113) 12076–12081. https://doi.org/10.1073/PNAS.1605086113/-/DCSUPPLEMENTAL.

[38] ICH Harmonized Tripartate Guideline Validation of Analytical Procedures: Text and Methodology Q2(R1), 1996. https://www.ich.org/page/quality-guidelines.

[39] SARS-CoV-2 Viral Mutations: Impact on COVID-19 Tests | FDA, (n.d.). https://www.fda.gov/medical-devices/coronavirus-covid-19-and-medical-devices/sars-cov-2-viral-mutations-impact-covid-19-tests (accessed February 22, 2022).

[40] J.J. Adashek, R. Kurzrock, Could mutations of SARS-CoV-2 suppress diagnostic detection?, Nat. Biotechnol. 2021 393. 39 (39) 274–275. https://doi.org/10.1038/s41587-021-00845-3.

[41] M.J. Jian, H.Y. Chung, C.K. Chang, J.C. Lin, K.M. Yeh, C.W. Chen, D.Y. Lin, F.Y. Chang, K.S. Hung, C.L. Perng, H.S. Shang, SARS-CoV-2 variants with T135I nucleocapsid mutations may affect antigen test performance, Int. J. Infect. Dis. 114 (114) 112–114. https://doi.org/10.1016/J.IJID.2021.11.006.

[42] L. Bourassa, G.A. Perchetti, Q. Phung, M.J. Lin, M.G. Mills, P. Roychoudhury, K.G. Harmon, J.C. Reed, A.L. Greninger, A SARS-CoV-2 Nucleocapsid Variant that Affects Antigen Test Performance, J. Clin. Virol. 141 (141) 104900. https://doi.org/10.1016/J.JCV.2021.104900.

[43] SARS-CoV-2 Variant Classifications and Definitions, (n.d.). https://www.cdc.gov/coronavirus/2019-ncov/variants/variant-classifications.html (accessed May 31, 2022).

[44] World Health Organization, Antigen-detection in the diagnosis of SARS-CoV-2 infection: interim guidance, 6 October 2021, 2021. https://www.who.int/publications/i/item/antigen-detection-in-the-diagnosis-of-sars-cov-2infection-using-rapid-immunoassays.

[45] M. Sidstedt, J. Hedman, E.L. Romsos, L. Waitara, L. Wadsö, C.R. Steffen, P.M. Vallone, P. Rådström, Inhibition mechanisms of hemoglobin, immunoglobulin G, and whole blood in digital and real-time PCR, Anal. Bioanal. Chem. 410 (410) 2569–2583. https://doi.org/10.1007/S00216-018-0931-Z/TABLES/5

