## Supplementary Materials for "Development of Q-LAAD, an allonamer-based antigen test for the rapid detection of SARS-CoV-2"

### Supplementary Material Captions

**Table S1: SARS-CoV-2 Antigen Diagnostic Tests with Emergency Use Authorization.**

List of diagnostic tests for the detection of SARS-CoV-2 with FDA Emergency Use Authorizations as of 31May22. Organized first by target antigen, then by company name in alphabetical order. A majority of the tests target the N-protein and are designed as lateral flow assays.

**Table S2: SARS-CoV-2 Variants and Current SARS-CoV-2 Interagency Group (SIG) Classification.**

List of SARS-CoV-2 Variants of Concern/Variants Being Monitored (both WHO label and PANGO lineage) with SIG classification as of 31May22. Includes location and time each variant was first detected.

**Table S3: Cross-Reactivity of High-Priority Pathogens Commonly Found in the Respiratory Tract Flora.**

High-priority pathogens that are commonly found in the respiratory tract flora were tested for cross-reactivity with the Q-LAAD assay for detection of SARS-CoV-2.

**Table S4: Interference of High-Priority Pathogens Commonly Found in the Respiratory Tract Flora.**

High-priority pathogens that are commonly found in the respiratory tract flora were tested for interference with the Q-LAAD assay for detection of SARS-CoV-2.

**Table S5: Interference of Endogenous Substances.**

Endogenous substances commonly present in clinical samples from the respiratory tract were tested for interference with the Q-LAAD assay for detection of SARS-CoV-2.

### Table S1: SARS-CoV-2 Antigen Diagnostic Tests with Emergency Use Authorization as of 22Sep2022[1]

| Company | Name of Diagnostic Test | Attributes[2] | Date EUA Issued | Antigen Targeted | Authorized Settings[3] |
|---|---|---|---|---|---|
| Celltrion USA, Inc. | Sampinute COVID-19 Antigen MIA | Magnetic Force-assisted Electrochemical Sandwich IA (MESIA), ST | 10/23/2020 | S-protein RBD | H, M |
| | Celltrion DiaTrust COVID-19 Ag Home Test | LF, VR, OTC, SS, MT | 10/21/2021 | N-protein S-protein RBD | Home, H, M, W |
| | Celltrion DiaTrust COVID-19 Ag Rapid Test | LF, VR, SS, MT | 04/16/2021 | | H, M, W |
| Abbott Diagnostics Scarborough, Inc. | BinaxNOW COVID-19 Ag Card | LF, VR, ST | 08/26/2020 | N-protein | H, M, W |
| | BinaxNOW COVID-19 Ag Card Home Test | LF, VR, OTC, TPS, SS, ST | 12/16/2020 | | Home, H, M, W |
| | BinaxNOW COVID-19 Ag 2 Card | LF, VR, Non-prescription Testing, SS, ST | 03/31/2021 | | H, M, W |
| | BinaxNOW COVID-19 Antigen Self Test | LF, VR, OTC, SS, ST | 03/31/2021 | | Home, H, M, W |
| Access Bio, Inc. | CareStart COVID-19 Antigen Home Test | LF, VR, OTC, SS, ST | 08/02/2021 | | Home, H, M, W |
| | CareStart COVID-19 Antigen test | LF, VR, SS, ST | 10/08/2020 | | H, M, W |
| ACON Laboratories, Inc | Flowflex COVID-19 Antigen Home Test | LF, VR, OTC, Screening, ST | 10/04/2021 | | Home, H, M, W |
| ANP Technologies, Inc | NIDS COVID-19 Antigen Rapid Test Kit | LF, VR, SS, ST | 09/24/2021 | | H, M, W |
| Becton, Dickinson and Company (BD) | BD Veritor System for Rapid Detection of SARS-CoV-2 | Chromatographic Digital IA, IR, SS, ST | 07/02/2020 | | H, M, W |
| | BD Veritor At-Home COVID-19 Test | LF, Digital Read, OTC, SS, ST | 8/24/2021 | | Home, H, M, W |
| DiaSorin, Inc. | LIAISON SARS-CoV-2 Ag | CLIA, ST | 03/26/2021 | | H, M |
| Ellume Limited | ellume.lab COVID Antigen Test | LF, Fluorescence, IR, ST | 07/08/2021 | | H, M, W |
| Ellume Limited | Ellume COVID-19 Home Test | LF, Fluorescence, IR, OTC, SS, ST | 12/15/2020 | | Home, H, M, W |
| Qorvo Biotechnologies, LLC. | Omnia SARS-CoV-2 Antigen Test | Bulk Acoustic Wave Biosensor, IR, ST | 04/13/2021 | | H, M |
| Genabio Diagnostics, Inc. | Genabio COVID-19 Rapid Self-Test Kit | LF, VR, OTC, SS, ST | 07/08/2022 | | Home, H, M, W |
| GenBody Inc. | GenBody COVID-19 Ag | LF, VR, SS, ST | 07/13/2021 | | H, M, W |
| iHealth Labs, Inc. | iHealth COVID-19 Antigen Rapid Test Pro | LF, VR, SS, ST | 01/14/2022 | | H, M, W |
| | iHealth COVID-19 Antigen Rapid Test | LF, VR, OTC, SS, ST | 11/05/2021 | | Home, H, M, W |
| InBios International Inc. | SCoV-2 Ag Detect Rapid Self-Test | LF, VR, OTC, SS, ST | 11/22/2021 | | Home, H, M, W |
| | SCoV-2 Ag Detect Rapid Test | LF, VR, SS, ST | 05/06/2021 | | H, M, W |
| Luminostics, Inc. | Clip COVID Rapid Antigen Test | LF immunoluminescent assay, IR, ST | 12/07/2020 | | H, M, W |
| LumiraDx UK Ltd. | LumiraDx SARS-CoV-2 Ag Test | Microfluidic Immunofluorescence Assay, IR, Screening, ST | 08/18/2020 | | H, M, W |
| Maxim Biomedical, Inc. | MaximBio ClearDetect COVID-19 Antigen Home Test | LF, VR, OTC, SS, ST | 01/19/2022 | | Home, H, M, W |
| Nano-Ditech Corp. | Nano-Check COVID-19 Antigen Test | LF, VR, SS, ST | 12/06/2021 | | H, M, W |
| Oceanit Foundry LLC | ASSURE-100 Rapid COVID-19 Test | LF, VR, ST | 2/28/2022 | | H, M, W |
| OraSure Technologies, Inc. | InteliSwab COVID-19 Rapid Test Rx | LF, VR, PHT, ST | 06/04/2021 | | Home, H, M, W |
| | InteliSwab COVID-19 Rapid Test | LF, VR, OTC, SS, ST | 06/04/2021 | | Home, H, M, W |
| | InteliSwab COVID-19 Rapid Test Pro | LF, VR, SS, ST | 06/04/2021 | | H, M, W |
| Ortho Clinical Diagnostics, Inc. | VITROS Immunodiagnostic Products SARS-CoV-2 Antigen Reagent Pack | Chemiluminescence IA, IR, ST | 01/11/2021 | | H, M |
| OSANG LLC | OHC COVID-19 Antigen Self Test | LF, VR, OTC, Serial Testing, ST | 04/06/2022 | | Home, H, M, W |
| PHASE Scientific International, Ltd. | INDICAID COVID-19 Rapid Antigen Test | LF, VR, SS, ST | 07/28/2021 | | H, M, W |
| | INDICAID COVID-19 Rapid Antigen At-Home Test | LF, VR, OTC, SS, ST | 3/16/2022 | | Home, H, M, W |
| QIAGEN GmbH | QIAreach SARS-CoV-2 Antigen | Digital LF, Fluorescence, IR, ST | 08/05/2021 | | H, M |
| Quidel Corporation | QuickVue SARS Antigen Test | LF, VR, SS, ST | 12/18/2020 | | H, M, W |
| | QuickVue At-Home OTC COVID-19 Test | LF, VR, OTC, SS, ST | 03/31/2021 | | Home, H, M, W |
| | Sofia SARS Antigen FIA | LF, Fluorescence, IR, SS, ST | 05/08/2020 | | H, M, W |
| | QuickVue At-Home COVID-19 Test | LF, VR, PHT, ST | 03/01/2021 | | Home, H, M, W |
| Salofa Oy | Sienna-Clarity COVID-19 Antigen Rapid Test Cassette | LF, VR, ST | 05/20/2021 | | H, M, W |
| SD Biosensor, Inc. | Pilot COVID-19 At-Home Test | LF, VR, OTC, SS, ST | 12/24/2021 | | Home, H, M, W |
| Siemens Healthcare Diagnostics, Inc. | Atellica IM SARS-CoV-2 Antigen | CLIA, Single Target | 3/11/2022 | | H, M |
| | ADVIA Centaur SARS-CoV-2 Antigen | CLIA, Single Target | 3/11/2022 | | H, M |
| Siemens Healthineers | CLINITEST Rapid COVID-19 Antigen Self-Test | LF, VR, OTC, SS, ST | 12/29/2021 | | Home, H, M, W |
| Watmind USA | Speedy Swab Rapid COVID-19 Antigen Self-Test | LF, VR, OTC, SS, ST | 07/08/2022 | | Home, H, M, W |
| Xiamen Boson Biotech Co., Ltd. | Rapid SARS-CoV-2 Antigen Test Card | LF, VR, OTC, Serial Testing, ST | 04/06/2022 | | Home, H, M, W |
| Xtrava Health | SPERA COVID-19 Ag Test | LF, VR, ST | 10/12/2021 | | H, M, W |
| Becton, Dickinson and Company (BD) | BD Veritor System for Rapid Detection of SARS-CoV-2 & Flu A+B | Chromatographic Digital IA, IR, Multi-analyte, ST | 03/24/2021 | N-protein Influenza A Influenza B | H, M, W |
| Princeton BioMeditech Corp. | Status COVID-19/Flu A&B | LF, VR, Multi-analyte, ST | 02/04/2021 | | H, M, W |
| Quidel Corporation | Sofia 2 Flu + SARS Antigen FIA | LF, Fluorescence, IR, Multi-Analyte, ST | 10/02/2020 | | H, M, W |

1 U.S. Food and Drug Administration. In Vitro Diagnostics EUAs - Antigen Diagnostic Tests for SARS-CoV-2. https://www.fda.gov/medical-devices/coronavirus-disease-2019-covid-19-emergency-use-authorizations-medical-devices/in-vitro-diagnostics-euas-antigen-diagnostic-tests-sars-cov-2. Accessed September 22, 2022.

2 Lateral Flow (LF), Visual Read (VR), Single Target (ST), Serial Screening (SS), Over-the-Counter Home Testing (OTC), Telehealth Proctor Supervised (TPS), Instrument Read (IR), Immunoassay (IA), Prescription Home Testing (PHT), Multiple Targets (MT)

3 H - Labs certified under Clinical Laboratory Improvement Amendments of 1988 (CLIA), 42 U.S.C. §263a, that meet requirements to perform high complexity tests.
M - Labs certified under CLIA, 42 U.S.C. §263a, that meet requirements to perform moderate complexity tests.
W - Patient care settings operating under a CLIA Certificate of Waiver.

**Table S2: SARS-CoV-2 Variants and Current SARS-CoV-2 Interagency Group (SIG) Classification[1]**

| WHO Label | PANGO Lineage | Location and Time First Detected | SIG Classification (as of 22Sep2022) |
|---|---|---|---|
| Alpha | B.1.1.7 and Q lineages | UK, Late 2020 | Variant Being Monitored |
| Beta | B.1.351 and descendent lineages | South Africa, Late 2020 | |
| Gamma | P.1 and descendent lineages | Brazil, Late 2020 | |
| Delta | B.1.617.2 and AY lineages | India, Late 2020 | |
| Omicron | B.1.1.529, BA.1, BA1.1, BA.2, BA.3, BA.4, and BA.5 lineages | South Africa, Nov 2021 | Variant of Concern |

[1] SARS-CoV-2 Variant Classifications and Definitions. https://www.cdc.gov/coronavirus/2019-ncov/variants/variant-classifications.html. Accessed September 22, 2022.

**Table S3: Cross-Reactivity of High-Priority Pathogens Commonly Found in the Respiratory Tract Flora**

| Organism | Strain | Source | Identifier | Concentration tested (CFU/mL and PFU/mL) | Number Detected (3 Replicates) | Cross-Reactivity |
|---|---|---|---|---|---|---|
| Human coronavirus | 229E | Isolate | ATCC Cat# VR-740 | $10^5$ | 0/3 | No |
| Human coronavirus | OC43 | Isolate | ATCC Cat# VR-1558 | $10^5$ | 0/3 | No |
| Rhinovirus | B632 | Isolate | ATCC Cat# VR-1645 | $10^5$ | 0/3 | No |
| *Haemophilus influenzae* | Rd [KW20] | Isolate | ATCC Cat# 51907 | $10^6$ | 0/3 | No |
| *Streptococcus pneumoniae* | 262 [CIP 104340] (Klein) Chester | Isolate | ATCC Cat# 49619 | $10^6$ | 0/3 | No |
| *Streptococcus pyogenes* | Bruno [CIP 104226] | Isolate | ATCC Cat# 19615 | $10^6$ | 0/3 | No |
| *Candida albicans* | CBS 562 [572, CCRC 20512, CECT 1002, DBVPG 6133, IFO 1385, IGC 3436, JCM 1542, NCYC 597, NRRL Y-12983] | Isolate | ATCC Cat# 18804 | $10^6$ | 0/3 | No |
| Pooled human nasal swab | N/A | N/A | N/A | N/A | 0/3 | No |
| Adenovirus 5 | Adenoid 75 | Isolate | ATCC Cat# VR-1516 | $10^5$ | 0/3 | No |
| Parainfluenza virus 3 | ATCC-2011-5 | Isolate | ATCC Cat# VR-1782 | $10^5$ | 0/3 | No |
| Parainfluenza virus 1 | C35 | Isolate | ATCC Cat# VR-94 | $10^5$ | 0/3 | No |
| Influenza A (H1N1) | A/WS/33 | Isolate | ATCC Cat# VR-1520 | $10^5$ | 0/3 | No |
| Influenza B | B/Florida/78/2015 | Isolate | ATCC Cat# VR-1931 | $10^5$ | 0/3 | No |
| Enterovirus | H | Isolate | ATCC Cat# VR-1432 | $10^5$ | 0/3 | No |
| Respiratory syncytial virus | Long | Isolate | ATCC Cat# VR-26 | $10^5$ | 0/3 | No |
| *Bordetella pertussis* | 18323 [NCTC 10739] | Isolate | ATCC Cat# 9797 | $10^6$ | 0/3 | No |
| *Mycoplasma pneumoniae* | Eaton Agent [NCTC 10119] | Isolate | ATCC Cat# 15531 | $10^6$ | 0/3 | No |
| *Staphylococcus aureus* | NCTC 8532 [IAM 12544, R. Hugh 2605] | Isolate | ATCC Cat# 12600 | $10^6$ | 0/3 | No |
| *Staphylococcus epidermidis* | FDA strain PCI 1200 | Isolate | ATCC Cat# CRM-12228 | $10^6$ | 0/3 | No |
| **Human coronavirus | NL63 (heat-inactivated) | Isolate | ZeptoMetrix Cat# 0810228CFHI | $1.70^5$ TCID 50/mL | 0/3 | No |
| Parainfluenza virus 2 | Greer | Isolate | ATCC Cat# VR-92 | $10^5$ | 0/3 | No |
| Parainfluenza virus 4b | 19503 | Isolate | BEI Cat# 3238 | $10^5$ | 0/3 | No |
| Influenza A H3N2 | A/Hong Kong/8/68 | Isolate | ATCC Cat# VR-1679 | $10^5$ | 0/3 | No |
| **MERS-CoV | EMC/2012 | Gamma-irradiated | BEI Cat# NR-50549 | $10^5$ | 0/3 | No |
| *SARS-CoV | Urbani | Gamma-irradiated | BEI Cat# NR-9323 | $10^5$ | 0/3 | No |
| Human Metapneumovirus | TN/83-1211 | Isolate | BEI Cat# NR-22227 | $10^5$ | 0/3 | No |
| *Chlamydiaceae, Chlamydia* | AR-39 | Isolate | ATCC Cat# 53592 | $10^6$ | 0/3 | No |
| *Legionella pneumophila* | Concord 3 [NCTC 11985] | Isolate | ATCC Cat# 33152 | $10^6$ | 0/3 | No |

### Table S4: Interference of High-Priority Pathogens Commonly Found in the Respiratory Tract Flora

| Organism (Identifier) | Pool | Concentration Tested (CFU/mL and PFU/mL) | Number Detected (3 Replicates) | Interference |
|---|---|---|---|---|
| *Mycoplasma pneumoniae (ATCC 15531)* | 1 | 1.00E+06 | 3/3 | No |
| *Streptococcus pneumoniae (ATCC 49619)* | | 1.00E+06 | | |
| *Haemophilus influenzae (ATCC 51907)* | | 1.00E+06 | | |
| Adenovirus 5 (ATCC VR-1516) | | 1.00E+05 | | |
| *Bordetella pertussis (ATCC 9797)* | | 1.00E+06 | | |
| *Streptococcus pyogenes ATCC (19615)* | 2 | 1.00E+06 | 3/3 | No |
| Rhinovirus (ATCC VR-1645) | | 1.00E+05 | | |
| Human coronavirus OC43 (ATCC VR-1558) | | 1.00E+05 | | |
| *Candida albicans (ATCC 18804)* | | 1.00E+06 | | |
| *Staphylococcus aureus (ATCC 12600)* | | 1.00E+06 | | |
| Human coronavirus 229E (ATCC VR-740) | 3 | 1.00E+05 | 3/3 | No |
| Respiratory syncytial virus (ATCC VR-26) | | 1.00E+05 | | |
| Parainfluenza virus 1 (ATCC VR-94) | | 1.00E+05 | | |
| *Staphylococcus epidermidis (ATCC CRM-12228)* | | 1.00E+06 | | |
| SARS-CoV (BEI NR-9323) | | 1.00E+05 | | |
| Parainfluenza virus 3 (ATCC VR-1782) | 4 | 1.00E+05 | 3/3 | No |
| Influenza A (H1N1) (ATCC VR-1520) | | 1.00E+05 | | |
| Influenza B (ATCC VR-1931) | | 1.00E+05 | | |
| Enterovirus (ATCC VR-1432) | | 1.00E+05 | | |
| MERS-CoV (BEI NR-50549) | | 1.00E+05 | | |
| Influenza A (H3N2) (ATCC VR-1679) | 5 | 1.00E+05 | 3/3 | No |
| *Chlamydiaceae (ATCC 53592)* | | 1.00E+06 | | |
| *Legionella pneumophila (ATCC 35096)* | | 1.00E+06 | | |
| Coronavirus NL63 (Zeptometrix 0810228CFHI) | | 1.00E+05 | | |
| Human Metapneumovirus (BEI NR-22227) | | 1.00E+05 | | |
| Human parainfluenza 2 (ATCC VR-92) | 6 | 1.00E+05 | 3/3 | No |
| Parainfluenza 4B (BEI NR-3238) | | 1.00E+05 | | |
| Pooled Nasal Wash | | N/A | | |

### Table S5: Cross-Reactivity and Interference of Endogenous Substances

| Potential interfering substances | Active Ingredient | (Recommended) Concentration Tested | Number Detected (3 Replicates) | Cross-Reactivity At Tested Concentration | Number Detected (3 Replicates) | Interference |
|---|---|---|---|---|---|---|
| Afrin | Oxymetazoline | 15% v/v | 0/3 | No | 3/3 | No |
| Azithromycin | Azithromycin | 250 ug/mL | 0/3 | No | 3/3 | No |
| Blood (Human) | Blood | 4% | 3/3 | Yes | 3/3 | No |
| Chloraseptic | Menthol | 1.5 mg/mL | 0/3 | No | 3/3 | No |
| Clathromycin | Clathromycin | 1 mg/mL | 0/3 | No | 3/3 | No |
| Flonase | Fluticasone Propionate | 5% v/v | 0/3 | No | 3/3 | No |
| Homeopathic | Alkalol | 1:10 dilution | 0/3 | No | 3/3 | No |
| Mucin | Purified mucin protein | 0.5% w/v | 2/3 | Yes | 3/3 | No |
| Mupirocin | Mupirocin | 10 mg/mL | 2/3 | Yes | 3/3 | No |
| Nasal Drops | Phenylephrine hydrochloride | 15% v/v | 1/3 | Yes | 3/3 | No |
| Nasal Spray | Cromolyn | 15% v/v | 0/3 | No | 3/3 | No |
| Nelimed | Hyaluronic Acid | 5% v/v | 0/3 | No | 3/3 | No |
| Phenol Throat Spray | Phenol | 15% v/v | 0/3 | No | 3/3 | No |
| Saline Nasal Spray | Saline | 5% v/v | 0/3 | No | 3/3 | No |
| Tamiflu | Oseltamivir | 5 mg/mL | 1/3 | Yes | 3/3 | No |
| Tobramycin | Tobramycin | 4 mg/mL | 0/3 | No | 3/3 | No |
| Zicam | Oxymetazoline hydrochloride | 5% v/v | 3/3 | Yes | 3/3 | No |
